# The relation of a blood DNA methylation score with six-year depression course and risk factors

**DOI:** 10.64898/2026.10.29.26366842

**Authors:** Dennis Klose, Yuri Milaneschi, Marco P M Boks, Nic J A van der Wee, Jeroen Lakerveld, Christiaan H Vinkers, Brenda W J H Penninx, Laura K M Han

## Abstract

**Background:** DNA methylation (DNAm) signatures of major depressive disorder (MDD) may be related to psychosocial, environmental, biological, and lifestyle risks. We tested a blood-based DNAm depression score (MD-MS) based on an existing EWAS (N = 24,754) in N = 731 individuals for predicting 8 clinical and 16 MDD risk factors and whether different MDD trajectories mapped to changes in MD-MS over six years.

**Methods:** We calculated the MD-MS based on the top 15 strongest, epigenome-wide significant depression associated sites from the largest EWAS to date in EPICv2-profiled blood samples from N = 366 NESDA participants at baseline and N = 365 at six-year follow-up (FU6). Sixty percent of participants obtained a lifetime MDD diagnosis before or during the study timeframe. Linear mixed effect models were used for all analyses.

**Results:** The MD-MS was elevated in lifetime MDD vs. controls (β=0.137, p=0.012). Lower social activity (β=0.051, p=0.043), CRP (β=0.068, p=0.006), triglycerides (β=0.055, p=0.036) and fasting glucose (β=0.059, p=0.025) showed positive associations with MD-MS, while higher neighborhood air pollution levels showed a negative association (β=-0.054, p=0.031). Adjustment for psychosocial variables (social activity, childhood trauma, partner status) attenuated the MD-MS association with lifetime MDD by 17.5% (β=0.114, p=0.051). Longitudinally, individuals with chronic MDD exhibited consistently higher MD-MS compared to controls (β = 0.295, p = 0.001).

**Conclusions:** The MD-MS captures an epigenetic signature of lifetime exposure to MDD. The significant influences of psychosocial and biological factors on this association, although of limited effect size, warrant further research on potentially actionable mechanisms against aberrant depression-related DNAm patterns. (249/250 words)

## Introduction

Major depressive disorder (MDD) is one of the most urgent health challenges worldwide, and its partially biological underpinnings are increasingly recognized (1, 2). While individual genetic variation influences personal MDD risk (3), epigenetic factors such as DNA methylation (DNAm) likely integrate MDD-associated risk exposures and behaviors into the organism with the potential to alter gene expression (4, 5). Such changes, measurable in blood immune cells, often correlate with DNAm levels in brain tissue (6). Independently of central nervous system mechanisms, blood immune cell DNAm changes may further predispose to cardiometabolic or other comorbidities in the context of MDD (7-10). They could occur before clinical MDD onset, as suggested in Shen et al., 2022 (11), or also arise as a consequence of MDD, potentially due to altered lifestyle behaviors or pharmacological treatment (12-15). Certain DNAm signatures may be indicative of disease outcomes and responsive to specific MDD treatments, but conclusive mechanistic understanding is currently lacking (16-19). Previous epidemiological studies have identified several psychosocial (20-22), environmental (23, 24), biological (25-27) and lifestyle factors (4, 12, 28, 29) which were associated to the presence of MDD, but few have specifically investigated whether they may be involved in shaping the epigenetic signature of MDD in blood (30).

As part of the Psychiatric Genomics Consortium (PGC), Shen et al., 2025 previously conducted a large epigenome-wide association study (EWAS) on major depression (MD), encompassing 24,754 blood samples of individuals of European ancestry from 18 cohorts (31). Of note, training samples included mostly self-reported MD but also clinically diagnosed MDD. From this work, an MD methylation score (MD-MS) was derived, shown to be elevated in individuals with a lifetime history of MD in an independent medical record dataset of N = 8,876 individuals. However, the psychosocial, environmental, biological and lifestyle factors driving elevated MD-MS in psychiatrically healthy and depressed populations, and how such epigenetic risk evolves longitudinally within individuals, remain unknown. Here, we leverage Illumina EPICv2 DNA methylation data from the Netherlands Study of Depression and Anxiety (NESDA (32)), involving a well-characterized sample including healthy controls and individuals with clinically diagnosed MDD, to examine the application of the MD-MS for 1.) detecting lifetime MDD, 2.) capturing a broad range of clinical, psychosocial, environmental, biological, and lifestyle exposures, and 3.) reflecting six-year MDD trajectories and thereby changes in symptomatology. Together, we aim to characterize the responsiveness of the MD-MS blood DNAm signature to MDD symptom changes and time-varying external exposures to better understand the importance of epigenetic factors to eventually improve MDD treatment practice.

## Methods

### Study design and participants

Data are from the Netherlands Study of Depression and Anxiety (NESDA), an ongoing multi-center, observational, naturalistic and longitudinal cohort study examining the course and consequences of depressive and anxiety disorders. A detailed description of the study rationale, design and methods is given previously (32). Briefly, 2,981 adults (18-65 years old) with or without a diagnosis of depression and/or anxiety disorder were recruited from community samples, primary care practices, and mental health organizations between 2004 and 2007. Detailed assessments were repeated after one, two, four, and six years of follow-up. Exclusion criteria were being diagnosed with a current clinically overt psychiatric disorder other than depression and anxiety as well as lacking proficiency in Dutch. Ethical approval was obtained centrally from the Ethical Review Board of the Vrije Universiteit Medical Centre (reference number 2003/183) in Amsterdam, the Netherlands, as well as from the Ethics Review Boards of the other participating research centers. All participants provided written informed consent.

### Selection criteria

To be included in this study, healthy controls from NESDA had to have an IDS-SR score < 14 at baseline. Further, only samples with sufficient volumes (> 30 µl) and concentration (> 50 ng/µl) of blood DNA were selected and participants without follow-up blood measurement at FU6 were omitted. Samples were further selected to have a balanced/normal distribution of age, sex, education and childhood trauma. Samples were also chosen to maximize the overlap with other available data modalities within NESDA. Some samples from the healthy control and remitted group were selected at random. Inclusion criteria for depression at baseline were (i) the presence of a DSM-IV MDD or dysthymia diagnosis (or both) in the past 6 months at baseline, established using the structure Composite International Diagnostic Interview AND (ii) confirmation of depressive symptoms in the month prior to baseline either by the CIDI or life chart interview AND (iii) availability of 2, 4 but in any case 6-year follow-up data on DSM-IV diagnosis and depressive symptoms (6-month recency).

### Disease course trajectories

For this subsample of N = 731, we selected persons based on MDD status as determined using the DSM-IV based CIDI psychiatric interview that was conducted at baseline and again after 2, 4 and 6 years of follow-up. Three specific MDD trajectories were deliberately represented among the chosen samples: New onset MDD (healthy at baseline, fulfilling MDD diagnosis during one of the NESDA FU waves over 6 years), remitted MDD (depressed at T0, but gets better at 2 years or 4 years and stays better at 6 years, stays better for at least 2 years) and chronic MDD (diagnosed with MDD at both baseline and follow-up, no remission in-between). The healthy trajectory refers to people with no clinical diagnosis of MDD from baseline to FU6 (40% of samples). Lifetime MDD refers to people who have experienced a clinical diagnosis of MDD at any point in their life, including those with a current diagnosis. Hence, this study can be described as an observational, trajectory-based, two-wave longitudinal design with repeated measures.

### Bisulfite conversion, DNA methylation measurement, quality control and data-preprocessing

Bisulfite conversion was performed at Erasmus MC Rotterdam. For this, 500 ng of genomic DNA underwent bisulfite conversion using the EZ-96 DNA Methylation Kit (Zymo Research Corporation, Irvine, CA, USA). Epigenotyping was performed on Illumina Infinium EPICv2 BeadChips (Illumina Inc., San Diego, CA, USA) on the iScanሢ platform in 2025. Baseline and FU6 data were preprocessed together in R. M-values were corrected for sample plate using the ComBat function from the sva R package to account for potential batch effects. For details see **Suppl. Information**.

### Calculation of major depression methylation score

EWAS summary statistics have been taken from https://doi.org/10.7488/ds/7929. The MD-MS is a weighted sum between EWAS-derived effect sizes and each of the epigenome-wide significant 15 CpG sites’ respective M value. For details see **Suppl. Information**.

### Clinical assessments

The MDD diagnoses and age of illness onset, and anxiety diagnoses were ascertained using structured CIDI interviews. MDD symptom severity was determined using the Inventory of Depressive Symptomatology (IDS-SR) (33). Anxiety symptoms were assessed using the Beck Anxiety Inventory (BAI-SR) (34). Antidepressant use was determined from unstructured interviews and ATC coded. Chronicity (% of past years spent depressed) was determined with a life chart interview (35).

### Other exposure variables

In total, 16 factors from four conceptual exposure domains (psychosocial, environmental, biological and lifestyle) were selected based on their relevance for MDD. Of note, we did not observe concerning levels of multicollinearity across our chosen determinants (max. Spearman’s *r* = 0.56 for neighborhood safety ∼ neighborhood air pollution, **Fig. S1**). For detailed information on psychosocial, environmental, biological, and lifestyle variables, which were available for both timepoints, please see **Suppl. Information**. For missingness in the chosen variables see **Table S1**.

### Statistical analysis

All statistical analyses have been conducted in RStudio v2024.12.0 (R v4.4.2) unless specifically stated otherwise. Model covariates were age, age^2^, sex and cell type proportions. For a full list of used R packages and detailed descriptions of Cohen’s d and AUC calculation and all model specifications see **Suppl. Information**.

## Results

### Major depression methylation score and lifetime MDD diagnosis

First, we calculated the major depression methylation score (MD-MS) in our cohort of controls and people with lifetime major depressive disorder (MDD) (**Table 1, Table S2**). While we use the term “lifetime MDD” throughout this article, many individuals in our lifetime MDD sample actually had a current MDD diagnosis (43% of samples), i.e. an active MDD episode at time of blood sampling, while a recent or past MDD diagnosis was less common and mostly refers to people with remitted MDD at FU6 (17% of samples). Among new onset samples, 74% showed MDD remission before FU6. The duration of current MDD episodes before baseline assessments was different for chronic MDD and remitted MDD groups at baseline (Wilcoxon rank sum test for percentage of time spent depressed in previous 4-5 years: median_chronic_ = 43%, median_remitted_ = 25%, p = 4.42×10^-6^). Longitudinal depressive symptom severity over the follow-up period is depicted in **Fig. S2**.

**Table 1.**
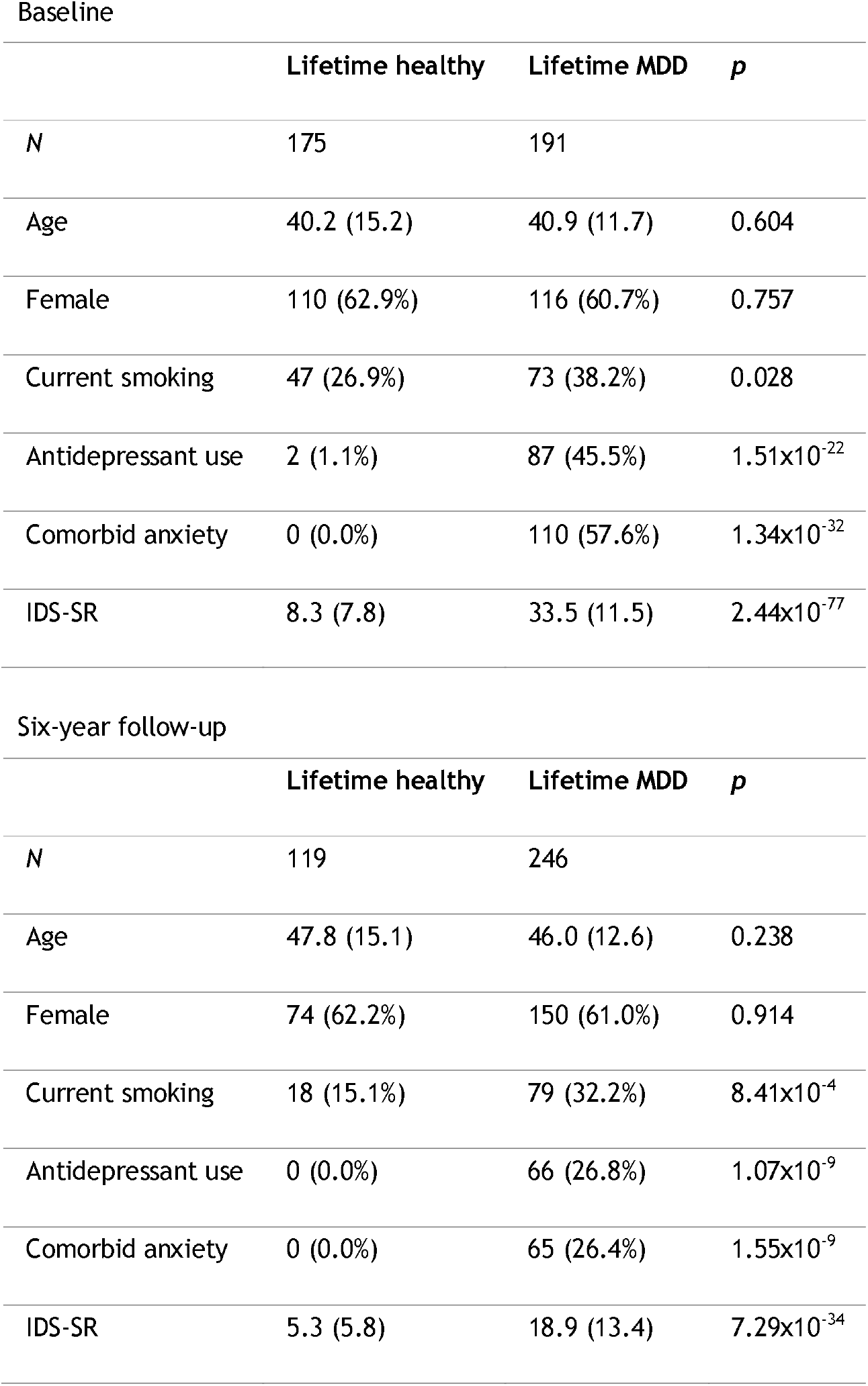
Cohort description with basic characteristics per timepoint. Age and IDS-SR: mean (SD). All other variables: *N* (%). P-values are derived from t-tests (continuous variables) and chi-square tests (categorical variables).

| Baseline |  |  |  |
|---|---|---|---|
|  | **Lifetime healthy** | **Lifetime MDD** | ***p*** |
| *N* | 175 | 191 |  |
| Age | 40.2 (15.2) | 40.9 (11.7) | 0.604 |
| Female | 110 (62.9%) | 116 (60.7%) | 0.757 |
| Current smoking | 47 (26.9%) | 73 (38.2%) | 0.028 |
| Antidepressant use | 2 (1.1%) | 87 (45.5%) | 1.51x10<sup>-22</sup> |
| Comorbid anxiety | 0 (0.0%) | 110 (57.6%) | 1.34x10<sup>-32</sup> |
| IDS-SR | 8.3 (7.8) | 33.5 (11.5) | 2.44x10<sup>-77</sup> |
| Six-year follow-up |  |  |  |
|  | **Lifetime healthy** | **Lifetime MDD** | ***p*** |
| *N* | 119 | 246 |  |
| Age | 47.8 (15.1) | 46.0 (12.6) | 0.238 |
| Female | 74 (62.2%) | 150 (61.0%) | 0.914 |
| Current smoking | 18 (15.1%) | 79 (32.2%) | 8.41x10<sup>-4</sup> |
| Antidepressant use | 0 (0.0%) | 66 (26.8%) | 1.07x10<sup>-9</sup> |
| Comorbid anxiety | 0 (0.0%) | 65 (26.4%) | 1.55x10<sup>-9</sup> |
| IDS-SR | 5.3 (5.8) | 18.9 (13.4) | 7.29x10<sup>-34</sup> |

The MD-MS was significantly increased in people with a lifetime MDD diagnosis compared to people who never had any MDD diagnosis (Cohen’s d = 0.21 [0.044, 0.374], β = 0.137 [0.03, 0.244], p = 0.012, **Figure 1A**). In a logistic regression model, we achieved discriminatory power of case versus control of AUC = 0.57 [95%CI: 0.53-0.61] (**Figure 1B**). Furthermore, we tested the association with overall symptom severity and antidepressant use. The association between current MD-MS and current symptom severity was not significant (β=0.009, p=0.716). The association between current antidepressant use and current MD-MS was comparable to that of lifetime MDD (Cohen’s d = 0.23 [0.039, 0.415], β = 0.149 [0.026, 0.272], p = 0.017) (**Table 2**). In a sensitivity analysis, we focused on subjects with current MDD and examined further clinical correlated exposures in **Table 2**. None of the clinical phenotypes were associated with MD-MS in this subsample, including depression severity (β = -0.027, p = 0.478) and current antidepressant use (β = 0.045, p = 0.553). No other clinical phenotype showed significant correlations to the MD-MS in neither full nor MDD subsample (MDD age of onset or chronicity, current anxiety diagnosis or symptoms, lifetime anxiety diagnosis). Altogether, we validated the PGC MD-MS within the independent NESDA samples and observed small group- and individual-level discriminatory effects with specificity for the lifetime MDD phenotype and current antidepressant use.

**Table 2.**
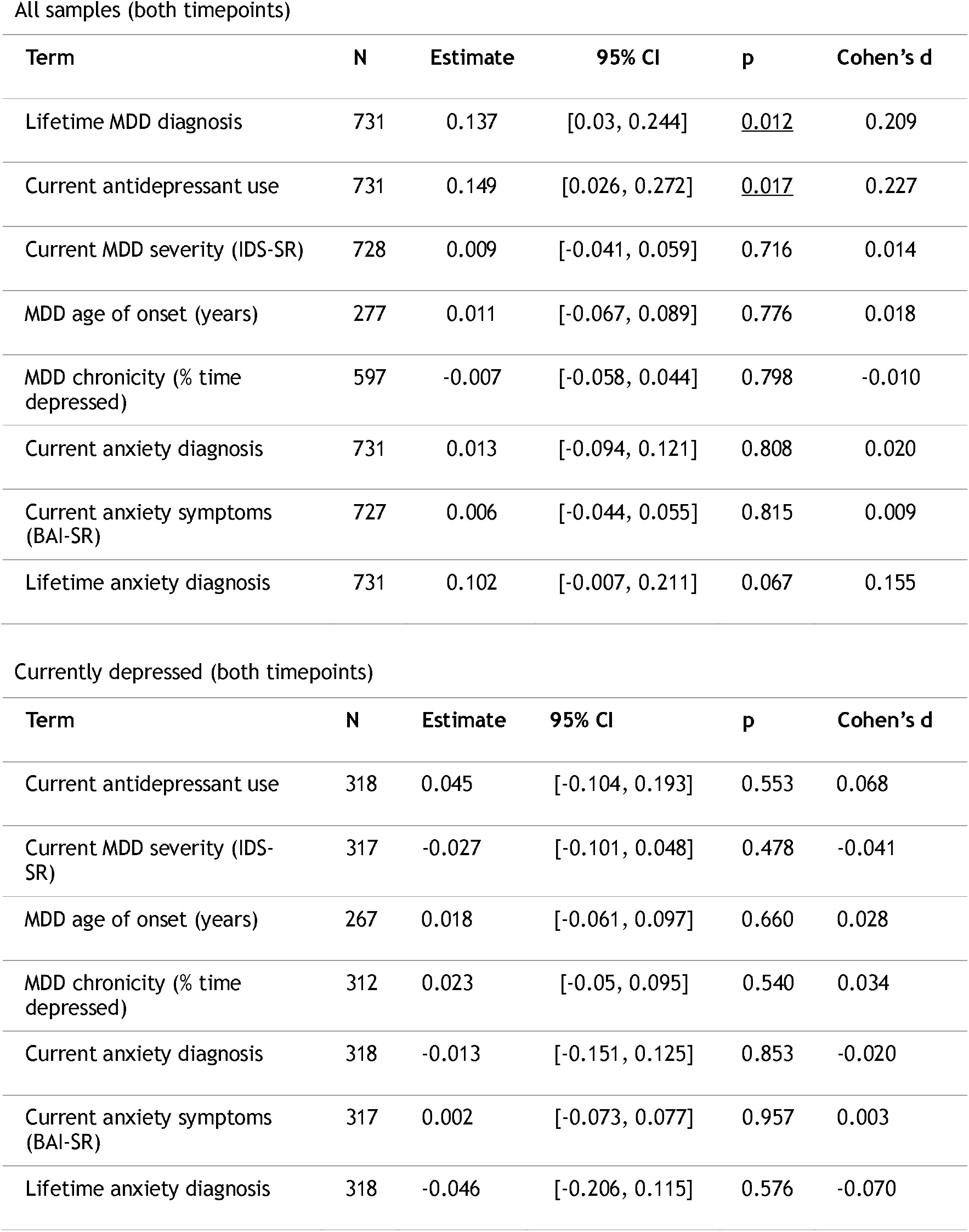
Results from cross-sectional (multi-level) LMEM analysis. Shown are estimates for other clinical, MDD-related phenotypes and their associations to MD-MS levels. Chronicity (% of time spent depressed) at baseline refers to past 4-5 years, whereas the last 2 years are the reference for FU6. First table has 10 more samples in age of onset than second table due to 3 new onset (baseline) and 7 remitted (FU6) samples with available data for age of MDD onset.

| All samples (both timepoints) |  |  |  |  |  |
| Term | N | Estimate | 95% CI | p | Cohen’s d |
|---|---|---|---|---|---|
| Lifetime MDD diagnosis | 731 | 0.137 | [0.03, 0.244] | <u>0.012</u> | 0.209 |
| Current antidepressant use | 731 | 0.149 | [0.026, 0.272] | <u>0.017</u> | 0.227 |
| Current MDD severity (IDS-SR) | 728 | 0.009 | [-0.041, 0.059] | 0.716 | 0.014 |
| MDD age of onset (years) | 277 | 0.011 | [-0.067, 0.089] | 0.776 | 0.018 |
| MDD chronicity (% time depressed) | 597 | -0.007 | [-0.058, 0.044] | 0.798 | -0.010 |
| Current anxiety diagnosis | 731 | 0.013 | [-0.094, 0.121] | 0.808 | 0.020 |
| Current anxiety symptoms (BAI-SR) | 727 | 0.006 | [-0.044, 0.055] | 0.815 | 0.009 |
| Lifetime anxiety diagnosis | 731 | 0.102 | [-0.007, 0.211] | 0.067 | 0.155 |
| Currently depressed (both timepoints) |  |  |  |  |  |
| Term | N | Estimate | 95% CI | p | Cohen’s d |
| Current antidepressant use | 318 | 0.045 | [-0.104, 0.193] | 0.553 | 0.068 |
| Current MDD severity (IDS-SR) | 317 | -0.027 | [-0.101, 0.048] | 0.478 | -0.041 |
| MDD age of onset (years) | 267 | 0.018 | [-0.061, 0.097] | 0.660 | 0.028 |
| MDD chronicity (% time depressed) | 312 | 0.023 | [-0.05, 0.095] | 0.540 | 0.034 |
| Current anxiety diagnosis | 318 | -0.013 | [-0.151, 0.125] | 0.853 | -0.020 |
| Current anxiety symptoms (BAI-SR) | 317 | 0.002 | [-0.073, 0.077] | 0.957 | 0.003 |
| Lifetime anxiety diagnosis | 318 | -0.046 | [-0.206, 0.115] | 0.576 | -0.070 |

**Figure 1.**
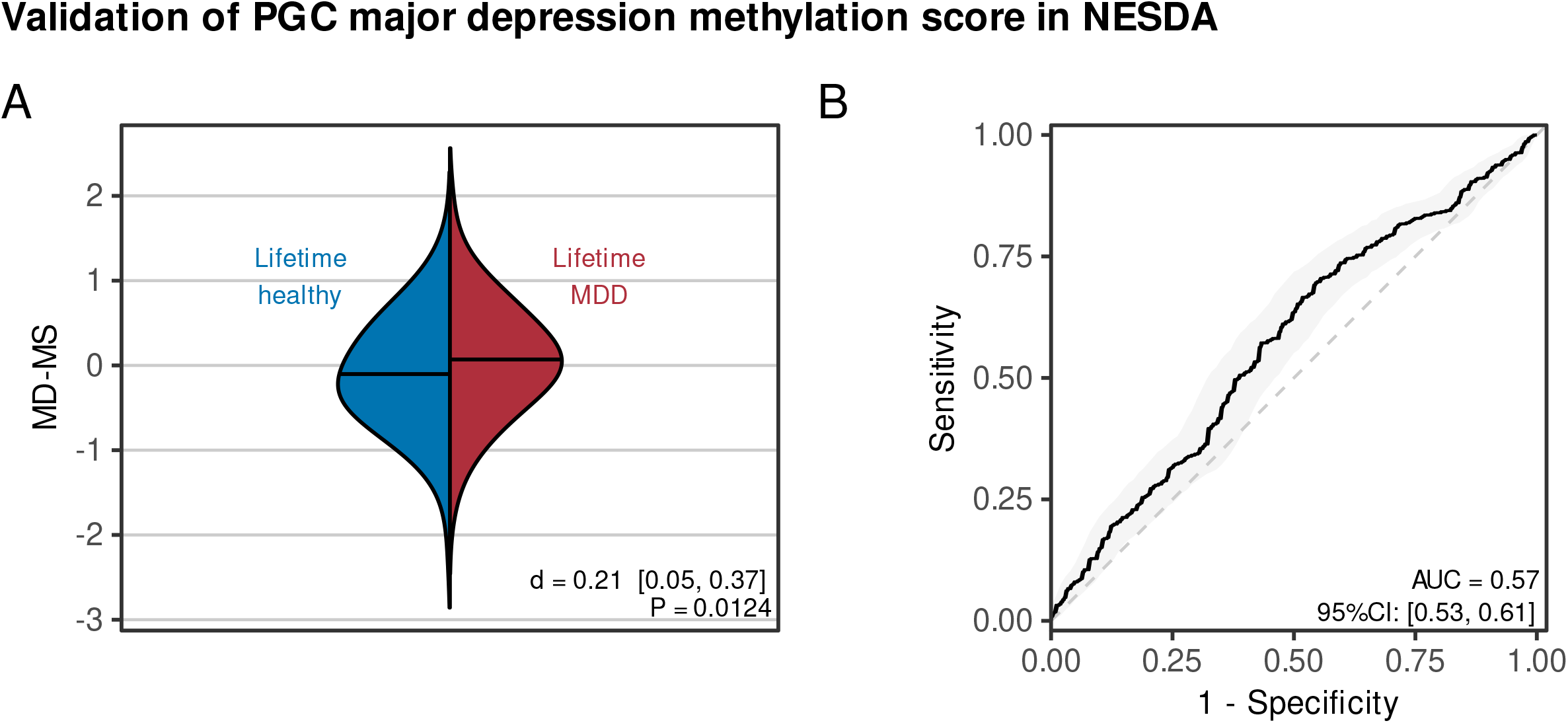
Validation of the PGC major depression methylation score in the independent NESDA sample. **A** Effect size (Cohen’s d) and t-test statistics for comparison of MD-MS in 731 samples from two timepoints from multi-level linear mixed effects model (random effects for paired participants). MD-MS is statistically significantly elevated in lifetime MDD participants compared to lifetime healthy controls. **B** Discriminative capability expressed as area under the receiving operator curve (AUC) of MD-MS between lifetime MDD and controls (MD-MS was residualized for fixed effects age, age^2,^ sex, cell type proportions, random effects are not residualized).

### Exposures underlying the association between MD-MS and a lifetime MDD diagnosis

Next, we aimed to understand whether the observed association between current MD-MS and lifetime MDD diagnosis could be partially explained by other factors, such as psychosocial, environmental, lifestyle or biological stressors which have been found to influence MDD. First, we assessed the univariate associations of these variables with the current MD-MS (N_max_=731; N may vary due to data missingness, **Table S3**; highest is 6% missingness in neighbourhood socioeconomic status). In the psychosocial domain, we observed a positive association between MD-MS and current low social activity (β = 0.051, p = 0.043, **Figure 2A**). In the environmental domain, we observed a significant negative association between MD-MS and higher current neighborhood air pollution (β = - 0.054, p = 0.031, **Figure 2B**). In the biological domain, we observed three significant positive associations between MD-MS and current CRP (β = 0.068, p = 0.006), triglycerides (β = 0.055, p = 0.036), and fasting glucose levels (β = 0.059, p = 0.025, **Figure 2C**). In the lifestyle domain, we observed no significant associations between MD-MS and the assessed MDD-risk factors (**Figure 2D**). We found in full-case analyses (N=654-722) that the association between current MD-MS and lifetime MDD was decreased by 17.5%, 9.4% and 4.9% when adjusting the regression coefficient of lifetime MDD for the psychosocial, environmental or biological domain variables, respectively (**Table S4**).

**Figure 2.**
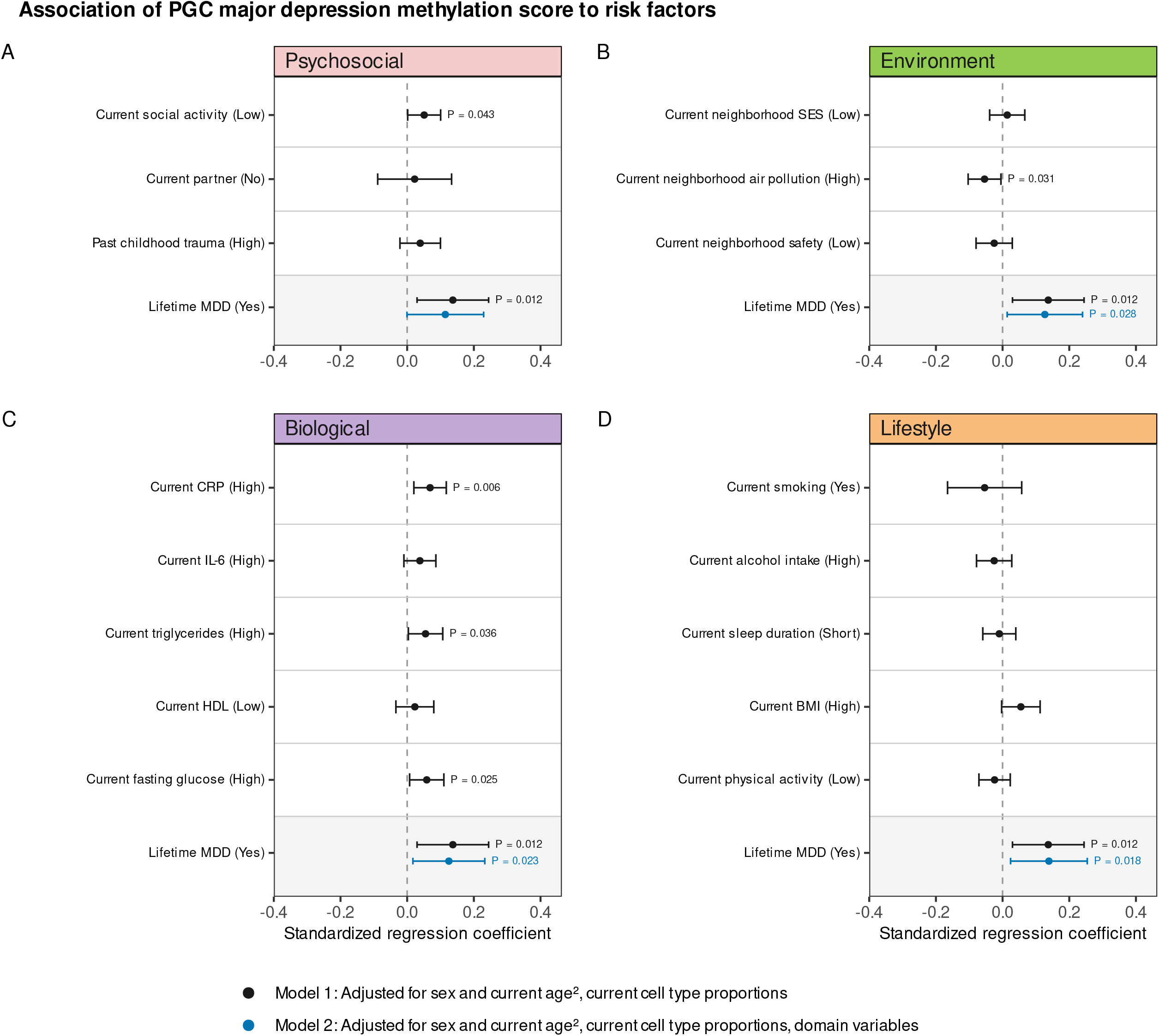
A mediation-like analysis for four domains and their role in the association of current MD-MS and lifetime MDD. **A** Psychosocial variables. Domain variables appear to partially mediate the association between lifetime MDD and MD-MS. Current social activity (lower) is statistically associated to higher MD-MS in univariate LMEM. **B** Environmental (GECCO) variables. Domain variables appear to lower the association between lifetime MDD and MD-MS but regression coefficient remains significant for lifetime MDD in multivariate model. Current neighborhood air pollution (higher) is statistically associated to lower MD-MS in univariate LMEM. **C** Biological variables. Domain variables appear to lower the association between lifetime MDD and MD-MS but regression coefficient remains significant for lifetime MDD in multivariate model. Current CRP (higher), current triglycerides (higher) and current fasting glucose in plasma (higher) are statistically associated to higher MD-MS in univariate LMEMs. **D** Lifestyle variables. Domain variables appear to strengthen the association between lifetime MDD and MD-MS. No factor is statistically associated to the MD-MS. All P-values are nominal.

We further performed sensitivity analysis using genetic principal components for the main analyses in 660 samples with genetic data, which suggested little influence of ancestry on MD-MS differences (**Fig. S3**). Interestingly, in a separate cross-sectional covariate-only model, we observed highly significant influences on MD-MS of the chosen covariates age (β = 12.28, p = 1.89×10^-28^), age^2^ (β = -4.59, p = 9.26×10^-7^), B-cells (β = -0.616, p = 6.32×10^-69^), CD4T cells (β = -0.564, p = 3.30×10^-61^), CD8T cells (β = -0.519, p = 6.96×10^-42^), NK cells (β = -0.211, p = 8.80×10^-8^), and neutrophils (β = 0.647, p = 9.87×10^-98^). The influences of sex (β = -0.085, p = 0.361) and AHRR cg-probe DNA methylation percentage as a proxy for smoke exposure (β = 0.094, p = 0.031) were small in comparison (**Fig. S4**). Since smoking was part of the main analyses shown in Figure 2, we decided to not model it as a covariate but as a variable of interest. To summarize, psychosocial factors, environment, and immuno-metabolic factors partially contributed to the association between current MD-MS and lifetime MDD.

### The six-year longitudinal trajectories of MD-MS

Finally, we modeled six-year longitudinal changes of the MD-MS, in relation to distinct disease courses and symptom severity changes. Across samples, we analyzed control participants, chronic MDD, remitted MDD, and new onset MDD (**Figure 3A, Table 3**). First, we saw that the overall effect of visit was significant (β = 0.193, p = 2.44×10^-6^, **Table S5**), indicating an increase in MD-MS in all groups over the course of six years. Further, there was a significant overall effect of chronic MDD course (β = 0.295, p = 0.001, **Table S5**). When adding an interaction in the model between visit and group, neither chronic (β = -0.047, p = 0.631), new onset (β = 0.028, p = 0.792) nor remitted MDD groups (β = 0.081, p = 0.343,) presented with different slopes (visit-by-group effects) compared to the control group (**Figure 3B, Table S5**). Pairwise comparisons per timepoint showed that estimated marginal means for MD-MS were consistently elevated in chronic MDD compared to controls at baseline (β = 0.319, p = 0.002) and FU6 (β = 0.272, p = 0.008). Chronic MDD had elevated MD-MS compared to remitted MDD at baseline (β = 0.222, p = 0.027), but not at FU6 (β = 0.094, p = 0.350). Remitted MDD did not differ from controls at baseline (β = 0.097, p = 0.270), but showed elevated MD-MS at FU6 (β = 0.178, p = 0.044) (**Figure 3B, Table S6**). When we decomposed IDS-SR for between- and within-person variance in a second longitudinal LMEM adjusted for baseline age^2^, sex and person-mean decomposed cell type proportions, we resolved significance for the (between) person-mean symptom severity (β = 0.081, p = 0.010), but not for longitudinal within-person changes (β = -0.026, p = 0.178, **Table S7**). In conclusion, these findings suggest that MD-MS differences at baseline remained stable, except for the remitted group. The MD-MS was consistently elevated in chronic disease course over six years. MD-MS increased over time in the whole study population.

**Table 3.**
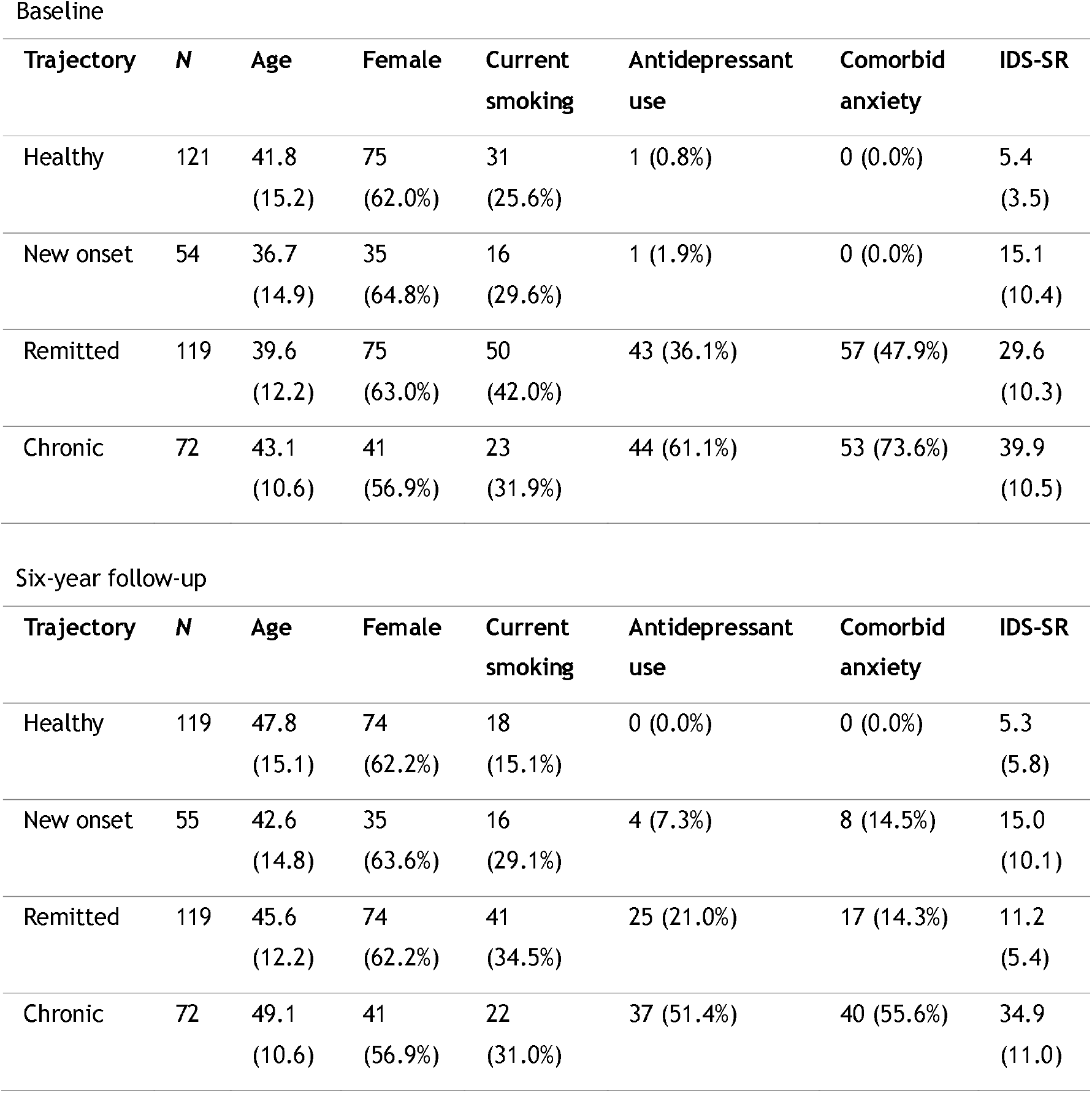
Cohort description per MDD trajectory. Age and IDS-SR: mean (SD). All other variables: *N* (%).

**Figure 3.**
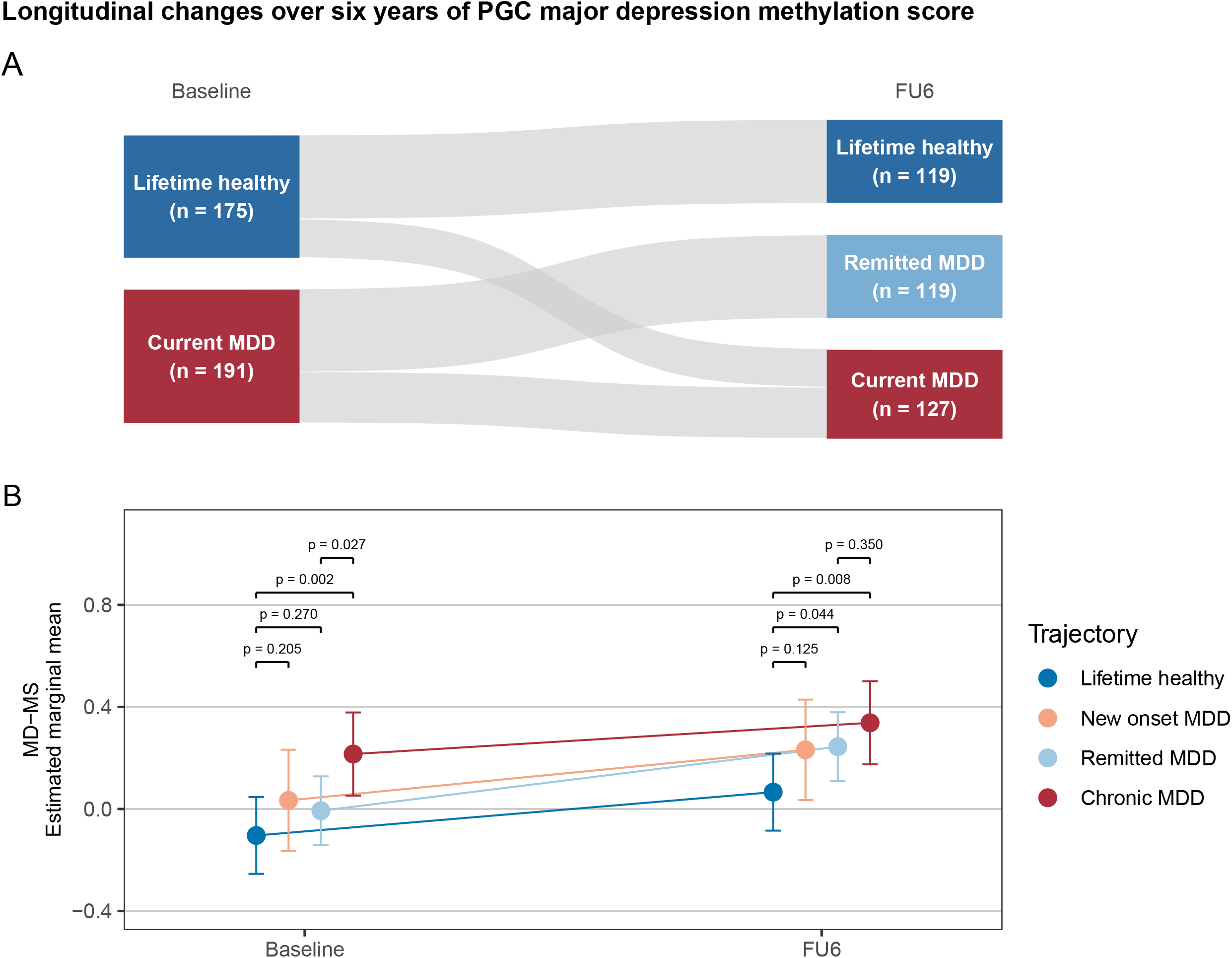
An analysis of six-year changes in the major depression methylation score in blood. **A** Sankey diagram for the four different MDD trajectories assessed in this study, including the lifetime healthy course. **B** Estimated marginal means from longitudinal LMEM for the four trajectories and contrasts between groups per timepoint. All P-values are nominal.

## Discussion

In this study, we validated the most recent PGC methylation score for major depression (MD-MS, (31)) in predicting lifetime major depressive disorder (MDD) in an independent dataset from the NESDA cohort including N = 731 repeated blood DNA methylation measurements at two timepoints six years apart. In particular, we were interested in other factors underlying MD-MS apart from lifetime MDD exposure, and the longitudinal 6-year change of the score in relation to different MDD disease courses. For this, we assessed 8 clinical and 16 exposome factors and their association to the MD-MS in controls and people with lifetime MDD. A higher MD-MS was associated with lower social activity and higher CRP, triglyceride and fasting glucose levels, but MD-MS was lower for high neighborhood air pollution. Longitudinally, the MD-MS increased over the course of six years, independent of MDD trajectory, although people with chronic MDD showed consistently elevated MD-MS over time compared to all other participants.

In our study, people with lifetime MDD had on average higher MD-MS than healthy controls, with low effect size and AUC (Cohen’s d = 0.21, AUC = 0.57) comparable to other cohorts using different MD-MS conceptualizations (4, 12, 31) or other biological markers associated with MDD (36-38). We achieved slightly higher effect sizes and AUC compared to the original publication by Shen et al. (Cohen’s d = 0.13, AUC = 0.53) in N = 8,876 individuals potentially due to different case definitions (31). In their testing sample, Shen et al. defined cases as people with ≥ 1 general practitioner (GP) diagnosis of major depression in their electronic health record; controls were defined as people without such entry. In contrast, cases in NESDA were people recruited from GP offices but also secondary mental health care institutions with a clinical diagnosis of MDD based on a psychiatric CIDI interview and included chronic high severity MDD cases, whereas controls reported an IDS score below 14 (i.e., low symptom severity) in addition to the absence of a lifetime MDD diagnosis.

The MD-MS was associated to lifetime MDD but not to current depressive symptom severity (IDS-SR). This might suggest that the epigenetic embedding of depression in the blood methylome is a reflection of stable underlying risk traits or a lasting mark of exposure to clinical depression and its lifestyle and behavioral correlates rather than a dynamic state marker of current disease activity. Clinically, the MD-MS for lifetime MDD was comparatively specific for the phenotype it was trained on, with only current antidepressant use showing similar effect sizes between cases and controls. In analyses focusing on MDD cases, clinical features such as current depressive symptoms, age of MDD onset, time spent depressed before assessments, current anxiety symptoms, current anxiety diagnosis and lifetime anxiety diagnosis did not show significant associations with the MD-MS.

When examining the impact of psychosocial, environmental, biological or lifestyle variables in the relationship between lifetime MDD and MD-MS, the psychosocial domain resulted in the highest reduction in association estimates (-17.5% vs. -9.4% for environmental and -4.9% for biological factors). However, it is important to mention that, unlike current social activity, current partner status and past childhood trauma were not significantly associated with MD-MS in separate models. Current social activity is a summed score of five self-report items from a questionnaire asking about the current frequency of social free time activities such as going to a bar, museum, restaurant, association, amusement park, cinema, club, or sports activity outside of home (from “almost never” to “a couple of times per week”). To rule out that alcohol intake or physical activity was the underlying factor beneath the association of MD-MS and social activity we calculated their correlations and found *r* of -0.24 and 0.19, respectively (Fig. S1), suggesting that it is more likely that decreased interpersonal contact contributed to the association between social activity and MD-MS. Social stress and its influence on DNA methylation changes have been studied extensively in rodents, primates and humans (39). For example, social isolation and living alone was associated with accelerated blood epigenetic age in men (40). Further, in a recent epigenome-wide association study in U.S. middle-aged twins, 25 suggestive CpG sites from blood were proposed to be linked to self-reported loneliness (41). Among the most studied physiological responses to social stress is increased cortisol production (42-44). However, studies have so far provided only early evidence regarding associations between cortisol exposure and blood DNA methylation (45, 46), leaving the biological mechanism underlying the association between MD-MS and social factors to be investigated in more detail.

It is noteworthy that CRP, triglyceride and fasting glucose levels showed significant positive associations to the MD-MS while these were uncorrelated to lifetime MDD itself (*r* between -0.02 and 0.06). This may warrant follow-up research on the potentially different interpretations of the MD-MS in non-depressed people and how it might relate to MDD vulnerability. Interestingly, we did not find an association for IL-6 and MD-MS as was earlier shown in Shen et al. using data from a community-based birth cohort (N = 875) (31).

Further, we observed that both current neighborhood air pollution and current smoking were associated to a lower MD-MS, a correlation which reached statistical significance for air pollution but not for smoking. The fact that both factors decreased the MD-MS, rather than increased it, as could be expected due to their status as risk factors for MDD (23, 24, 47-49), is challenging to explain mechanistically and may simply be a spurious association observed due to selection (collider) bias in our sample with enrichment of MDD cases (50).

To our knowledge, this is the first study to demonstrate elevated MD-MS scores in individuals with a clinical diagnosis of MDD compared with healthy controls and to examine these differences longitudinally across two time points six years apart in a large well-characterized clinical sample. This study is therefore also the first comprehensive investigation of 24 risk factors associated with MDD contributing to the MD-MS. Despite these important strengths, several limitations are noted. First, we analyze MDD-related DNAm signatures in whole blood, whose concordance with brain DNAm is debated and may vary between brain regions (51, 52). Second, due to the restriction of our analysis to the top 15 CpGs with epigenome-wide significance, we may have optimized the detection of lifetime MDD versus healthy samples at the cost of detecting associations to lifestyle or environmental factors, which may affect CpG sites at less stringent P-value thresholds. Third, we cannot exclude the influence of other sources of heterogeneity, including differences in sample characteristics (e.g., ethnicity) and technical or methodological factors (e.g., sample freezer time), which may have affected our findings and their generalizability. Finally, we did not correct the reported P-values for multiple testing and therefore underline that the presented results should be seen as descriptive, exploratory and hypothesis-generating rather than providing mechanistic explanations. It would be of interest to validate these findings in a targeted way in larger cohorts with comparable data (55).

Further research is needed to better understand whether people with MDD scoring high on MD-MS show different treatment responses or outcomes than people with “low DNAm burden”. Finally, we assume that DNAm marks linked to MDD in blood could provide further explanations for comorbidities of MDD with metabolic or cardiovascular health conditions (7), but this needs to be tested in larger cohorts with an enrichment for such conditions. Ultimately, the potential for reversibility of MDD-related DNAm marks and how this affects physical and mental health should be assessed to better understand the clinical relevance of blood DNAm in MDD.

## Conclusions

Overall, our study confirms that blood DNAm signatures are a promising molecular readout of the biological embedding of MDD and its associated risk factors. However, its exact place in the etiology of the disorder remains unclear. Our results suggest that social activity and immuno-metabolic mechanisms may partially explain the epigenetic signature of lifetime MDD and could therefore be considered potential targets for its modulation. High DNAm-burden for MDD may further be indicative of chronic MDD courses, although this observation warrants further replication in people with MDD with similar symptom severity and chronicity. Finally, DNAm-burden for MDD appears to rise over time in both healthy and depressed people, putting forward the potential value of further research of aging mechanisms in the context of MDD.

## Data Availability

Code is available from Zenodo at https://doi.org/10.5281/zenodo.23495761. The data used to support the findings of this study are available upon reasonable request from NESDA, Amsterdam. Information on how to request the study data, including the data sharing policy, can be found at https://www.nesda.nl/nesda-english/. Environmental exposure data can be obtained through www.gecco.nl.

## Acknowledgements

First, we thank all NESDA participants for their valuable time and cooperation, without them, this study would not have been possible. The infrastructure for the NESDA study (https://www.nesda.nl/) is funded through the Geestkracht program of the Netherlands Organisation for Health Research and Development (ZonMw, grant number 10-000-1002) and financial contributions by participating universities and mental health care organizations (VU University Medical Center, GGZ inGeest, Leiden University Medical Center, Leiden University, GGZ Rivierduinen, University Medical Center Groningen, University of Groningen, Lentis, GGZ Friesland, GGZ Drenthe, Rob Giel Onderzoekscentrum). DK and BWJHP are supported by the research project Stress in Action – Innovation in the understanding and application of daily-life stress. which is financially supported by the Dutch Research Council and the Dutch Ministry of Education, Culture and Science (NWO gravitation Grant No. 024.005.010). YM has received funding from Amsterdam UMC (Starter Grant Ronde 2), Amsterdam Neuroscience (PoC funding 2024–2026) and ImmunoMIND, funded by UK Research and Innovation as part of the UK national Mental Health Platform. LKMH was funded by the Veni award (Grant No. 09150162210201) from the Dutch Research Council (NWO). This work was supported in part by a Vidi grant from the Dutch Research Council (NWO; grant number 09150171910042) awarded to CHV. Additionally, we thank the Advanced Compute and Data Core (ACDC) of Amsterdam UMC for providing the high-performance computing resource that supported this research. Geo-data were collected as part of the Geoscience and Health Cohort Consortium (GECCO), which was financially supported by the Netherlands Organization for Scientific Research (NWO), the Netherlands Organization for Health Research and Development (ZonMw), and Amsterdam UMC. More information on GECCO can be found on www.gecco.nl. We thank Alfred Wagtendonk and Tim Zandbelt for their assistance with GECCO data management. Generative AI (Anthropic Claude Sonnet 4.6-5.5, Opus 4.8-5.5) has been used to support coding (analysis/plotting) and draft the paragraph for supplemental information about statistics. The AI was primarily used to assist with understanding concepts, generating code, debugging, and improving code structure and readability. Prompts: Targeted prompts describing the problem were formulated and specified desired functionality or response to error messages. Prompts were iteratively refined to improve the relevance and accuracy of the code. Draft text for statistics in supplemental methods was refined and modified to own understanding. Settings: Default AI settings were used without customization. No sensitive or confidential data was shared with the AI tool. All outputs were treated as suggestions rather than final solutions. Methodology: AI-generated code and explanations were critically evaluated before use. It was reviewed, tested, and modified to ensure overall correctness and its alignment with project requirements. The final implementation reflects our own understanding and validation of the solutions.

## Conflict of interest

The authors declare no conflict of interest.

## Author contributions

DK: Conceptualization, Methodology, Software, Validation, Formal analysis, Writing - Original Draft, Visualization; LKMH: Conceptualization, Methodology, Investigation, Resources, Data Curation, Writing - Review & Editing, Supervision, Project administration; MB: Writing - Review & Editing; NJAvdW: Writing - Review & Editing; JL: Data Curation, Writing - Review & Editing; CHV: Project administration, Writing - Review & Editing, Funding acquisition; YM: Methodology, Writing - Review & Editing, Supervision; BWJHP: Methodology, Supervision, Project administration, Writing - Review & Editing, Funding acquisition.

## Notes

### Competing Interest Statement

The authors have declared no competing interest.

### Author Declarations

Ethical approval was obtained centrally from the Ethical Review Board of the Vrije Universiteit Medical Centre (reference number 2003/183) in Amsterdam, the Netherlands, as well as from the Ethics Review Boards of the other participating research centers. All participants provided written informed consent.

