## Supplementary figures for "The relation of a blood DNA methylation score with six-year depression course and risk factors"

### Slide 1
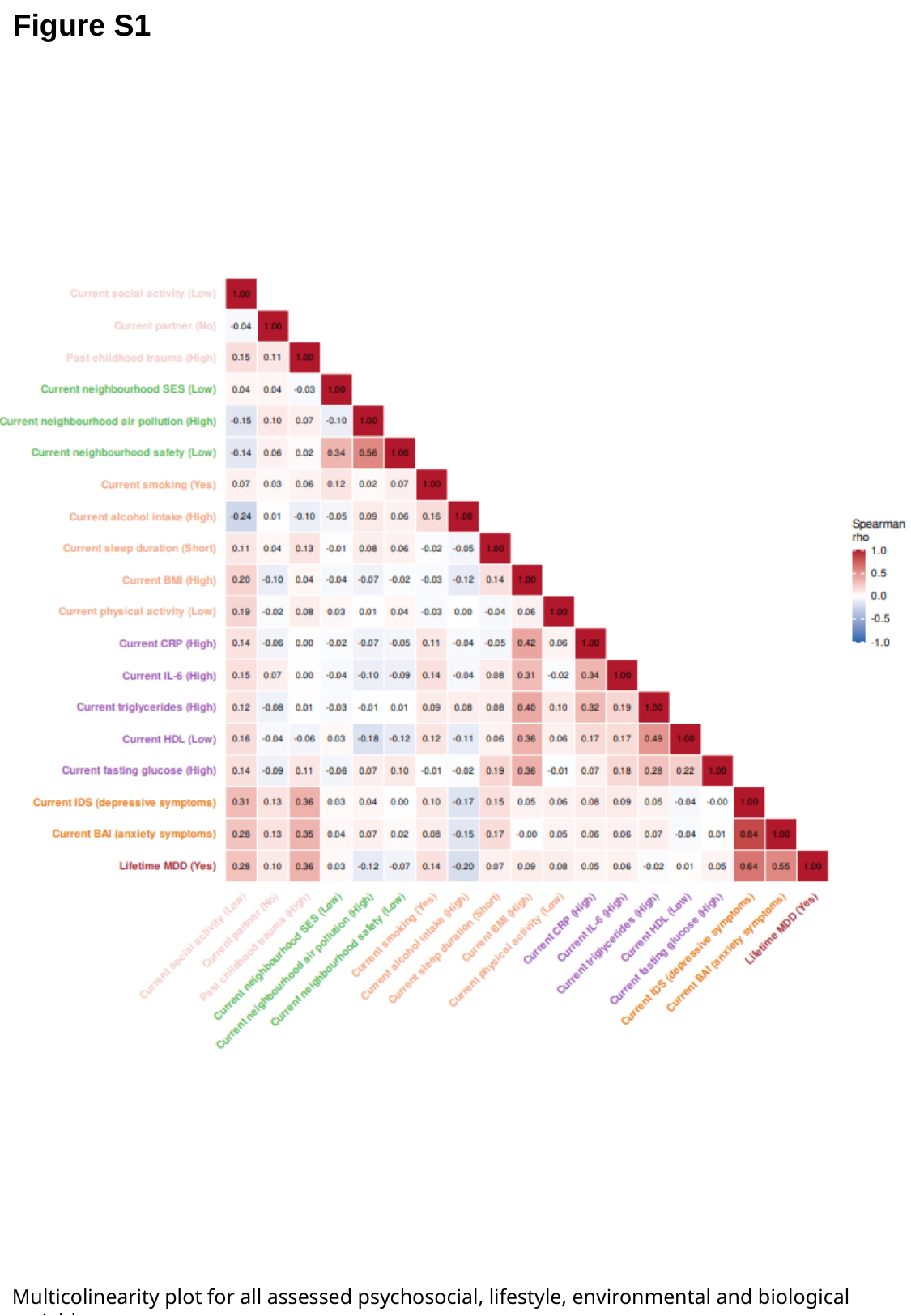

Figure S1
Multicolinearity plot for all assessed psychosocial, lifestyle, environmental and biological variables.

### Slide 2
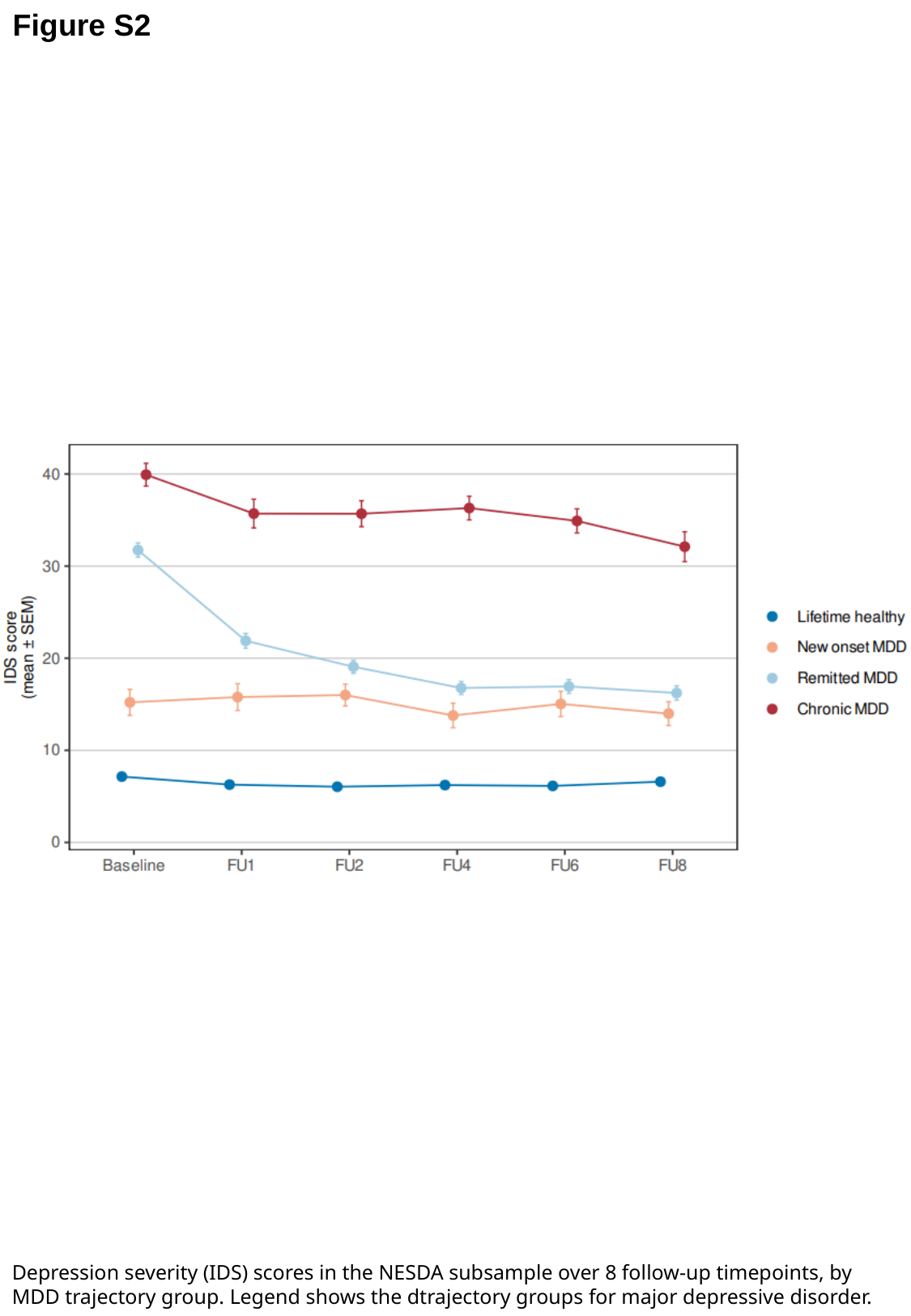

Figure S2
Depression severity (IDS) scores in the NESDA subsample over 8 follow-up timepoints, by MDD trajectory group. Legend shows the dtrajectory groups for major depressive disorder.

### Slide 3
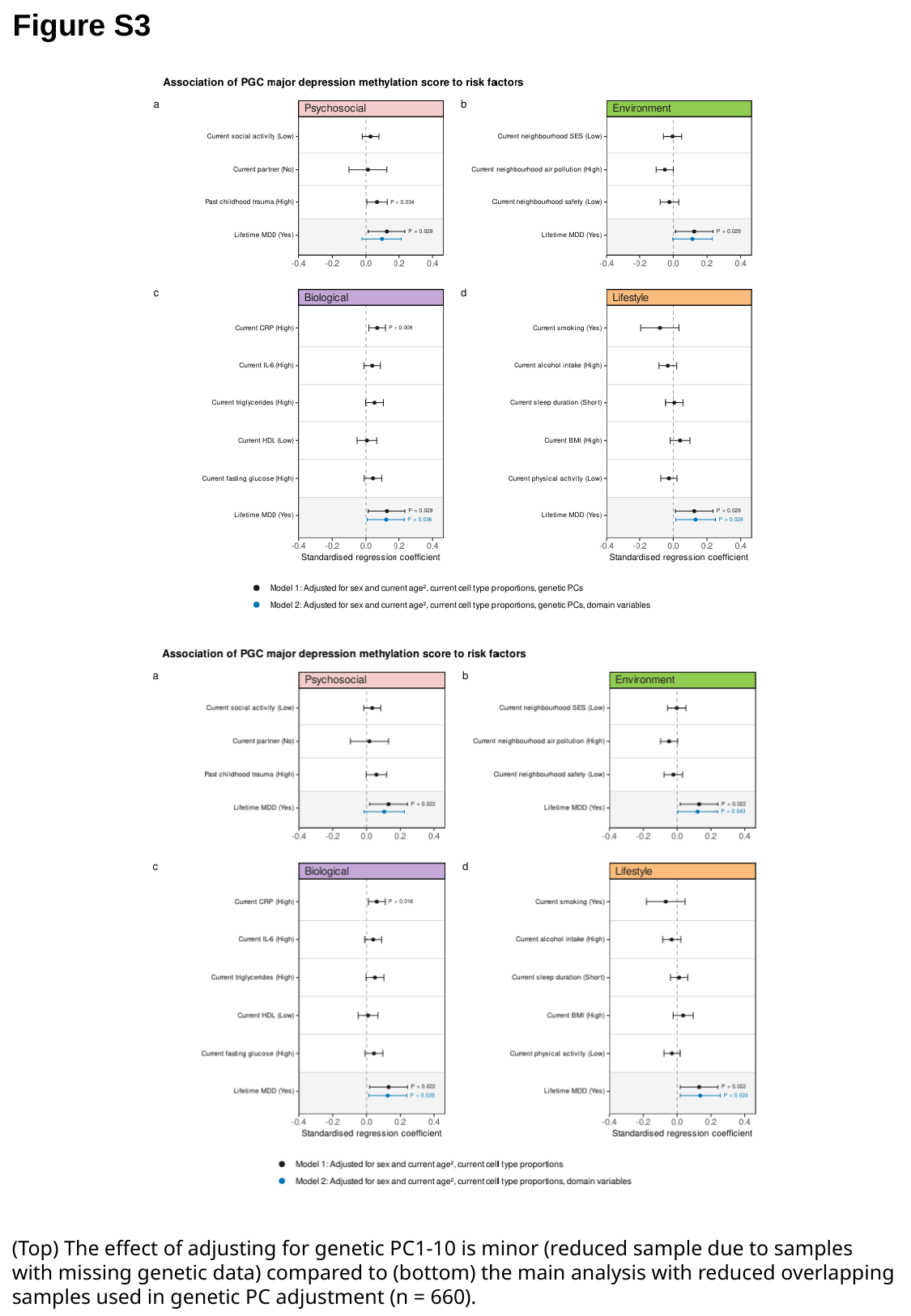

Figure S3
(Top) The effect of adjusting for genetic PC1-10 is minor (reduced sample due to samples with missing genetic data) compared to (bottom) the main analysis with reduced overlapping samples used in genetic PC adjustment (n = 660).

### Slide 4
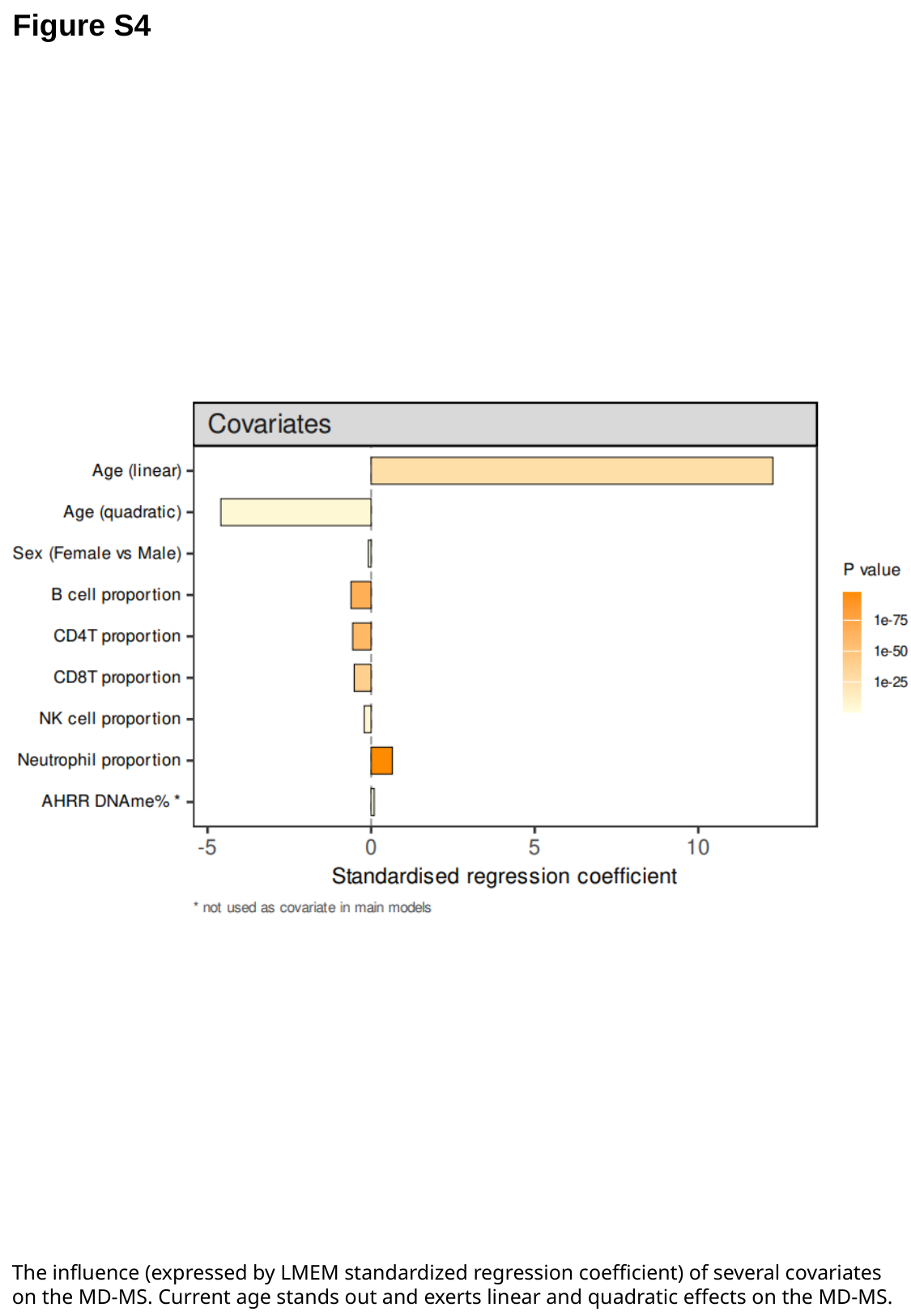

Figure S4
The influence (expressed by LMEM standardized regression coefficient) of several covariates on the MD-MS. Current age stands out and exerts linear and quadratic effects on the MD-MS.
