## Supplementary information for "The relation of a blood DNA methylation score with six-year depression course and risk factors"

*Exposures*

In brief, for the psychosocial domain, we assessed current social activity, current partner status and past childhood traumatic experiences through self-report. For the environmental domain we assessed current neighborhood socioeconomic status (SES), current neighborhood air pollution and current neighborhood safety - indicators that were provided by the Netherlands Institute of Social Research (SES), the Institute for Risk Assessment Science and the Institute for Public Health and the Environment (air pollution) and the Netherlands Ministry of the Interior and Kingdom Relations (neighborhood safety) and linked to NESDA participants through current postal codes in the Dutch PC4 format. In the biological domain, we assessed current statuses of C-Reactive Protein (mg/l, CRP), Interleukin-6 (pg/ml, IL-6), triglycerides (mmol/l), HDL-cholesterol (mmol/l) and fasting glucose (mmol/l) through laboratory assays. In the lifestyle domain, we assessed current statuses of smoking (current vs non-current), alcohol intake (drinks per week), sleep duration (hours), BMI (kg/m^2^) and physical activity (MET min) through self-report questionnaires.

Psychosocial: The *social activity score* is a summed score of five items from a questionnaire asking about the current frequency of social free time activities such as going to a bar, museum, restaurant, association, amusement park, cinema, club or sports activity outside of home. *Partner status* was assessed via a basic intake questionnaire at study begin. The reported *childhood trauma score* was assessed using the Childhood Trauma Questionnaire from the NEMESIS study and asked about traumatic experiences such as parental divorce, juvenile prison detention, foster family or child home placement, emotional neglect and psychological, physical or sexual abuse in the first 16 years of life.

Lifestyle: When categorical, *smoking status* was determined based on data originating from the Fagerström Questionnaire and is expressed as never smoker, former smoker and current smoker. When used as a continuous variable in line with Shen et al. (2025) for sensitivity analyses, the (1 – DNAme%) of cg05575921 (AHRR gene) was used as a smoke exposure proxy, which has been shown to be more reliable than self-report. *Alcohol intake* was calculated based on data originating from the AUDIT Questionnaire and is expressed as drinks per week. Sleep duration was derived from the Insomnia Rating Scale questionnaire[1]. *Physical activity* was assessed using the International Physical Activity Questionnaire and is expressed as MET-minutes per week, capturing sports activity, movement and commuting[2]. *BMI* was determined as current body weight [kg] /height^2^ [m].

Environment: Environmental data has been provided by the GECCO consortium of Amsterdam UMC[3, 4]. Baseline annual average air pollution concentra­tions at the subjects’ home addresses were estimated by land-use regression models for the year 2009 by the Institute for Risk Assessment Sciences as part of the European Study of Cohorts for Air Pollution Effects (ESCAPE-project), as described elsewhere[5, 6]. This study uses data of the annual average concentrations of the mean blackness of PM_2.5_ filters, which is a proxy for elemental carbon (EC), the dominant light absorbing substance. For FU6 air pollution exposures we used comparable EC data delivered by the Institute for Public Health and the Environment (RIVM), referring to large scaled concentration in µg/m³ maps (1x1 km resolution) with annual average outdoor concentration. Data are based on a combination of model calculations and measurements from official measurement locations. The socioeconomic status scores (SES) for baseline and FU6 were obtained from the Netherlands Institute of Social Research (SCP). The SES score is based on three dimension scores: education, income and position in the labor market. A principal component analyses defined the SES score based on mean income, percentage low incomes, percentage of low educated residents and percentage of unemployed residents. The neighbourhood social safety score for baseline and FU6 has been measured as part of the assessment of neighbourhood livability by the Netherlands Ministry of the Interior and Kingdom Relations[7] (Leefbarometer). The safety score (range: ­‑50 to 50) is based on indicators such as annoyance (of drug abuse, loitering youth, destruction, rubbish and daubing/graffiti) and number of crimes.

Biological: For inflammation marker assays fasting blood samples of NESDA participants were obtained in the morning around 8 am and kept frozen at -80°C. CRP was assayed at the Clinical Chemistry department of the VU University Medical Center. High-sensitivity plasma levels of CRP were measured in duplicate by an in-house ELISA based on purified protein and polyclonal anti‑CRP antibodies (Dako, Glostrup, Denmark). The CRP assay was standardized against the CRM 470 reference agent. IL-6 was assayed at the Clinical Chemistry department of the VU University Medical Center. Plasma IL-6 levels were measured in duplicate by a high sensitivity enzyme-linked immunosorbent assay (PeliKine Compact^TM^ ELISA, Sanquin, Amsterdam, The Netherlands). The IL-6 assay was standardized against a recombinant human IL-6 standard. In this study, CRP and IL-6 levels were ln-transformed for linear modelling. Fasting triglycerides, HDL cholesterol and glucose levels were determined using standard laboratory methods. Use of lipid modifying, antihypertensive or anti-diabetic medication in the past month was registered by drug container inspection and ATC coded and has been regressed out from the triglyceride, HDL, blood pressure and glucose variables.

*Data preprocessing*

Illumina EPICv2 preprocessing pipeline: In total, 735 samples were sequenced, but 4 samples were excluded after sequencing quality control, leaving 366 baseline and 365 FU6 samples. Samples from the same individual from both timepoints were assayed on the same array and randomized across plates. Randomization across sample plates and arrays was performed based on MDD course trajectory, age, sex and smoking status. Sample- and probe-based QC was performed by Erasmus MC Rotterdam using R version 4.3.0 and they involved the use of minfi, meffil and ewastools R packages. Basic quality control metrics were derived as described in the [BeadArray Controls Reporter Software Guide](https://support.illumina.com/content/dam/illumina-support/documents/documentation/chemistry_documentation/infinium_assays/infinium_hd_methylation/beadarray-controls-reporter-user-guide-1000000004009-00.pdf) by Illumina. Epigenotyped samples were SNP-matched to previously available genetic data from Affymetrix chips to confirm sample identity.

For the import of sample metadata, the minfi read.metharray.sheet function was used. For the import of raw .idat files, the minfi read.metharray.exp function was used, with ‘extended’ set to true. After import, four samples were immediately excluded from further preprocessing, one due to suboptimal sample detection *P*-value levels, two due to BeadArray Controls Reporter metrics and one due to a predicted sex mismatch. To confirm the absence of switched sample, we matched the SNP data provided from the Illumina EPICv2 array with Affymetrix-based genetic data from NESDA and compared the genotype (see next section in Suppl. Information). Furthermore, before any preprocessing, all EPICv2 “nv”-probes were removed, as those denote cancer-risk mutations and do not measure DNA methylation levels. Next, we used the minfi preprocessFunnorm function for functional between-array normalization, while accounting for sample sex. Background and dye-bias correction were performed, ‘ratioConvert’ was set to false. Then, potentially SNP-biased probes were removed using the minfi functions addSnpInfo and dropLociWithSnps, with ‘snps’ and ‘maf’ set to ‘CpG’ and ‘0’, respectively. After this, all probes with detection *P*-value < 0.05 and bead count < 3 were set to missing. Further, all probes which have previously been reported to be cross-reactive, flagged or flawed by mapping inaccuracies were set to missing. The resulting beta matrix was concatenated to a placeholder matrix for 450k/EPIC850k probes which were missing on the EPICv2 but necessary for MD-MS calculation. All missing values were then imputed using the OSMI method[8], taking the respective DNA methylation level of the nearest measured probe for the missing probe. The resulting matrix was then collapsed using the sesame betasCollapseToPfx function, to average the beta value of EPICv2 technical replicate probes. Finally, ComBat from the sva package was used to correct M values for any remaining sample plate effect. M values were converted back to betas using the lumi m2beta() function.

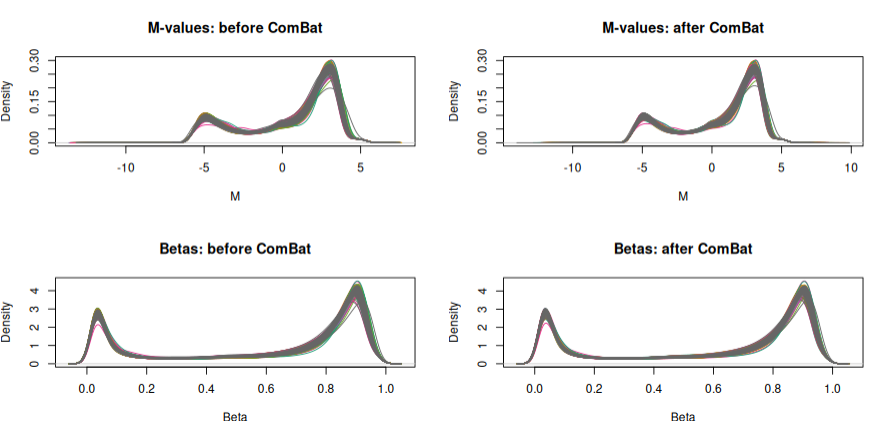

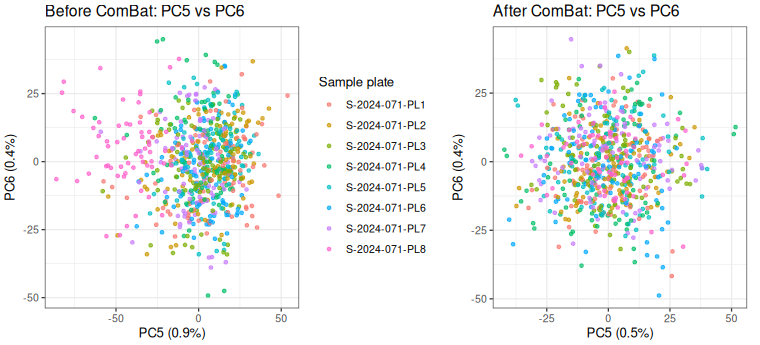

*Samples (both waves together) colored by sample plate. ComBat on M values removed residual sample plate effects, visualized in PC5 and PC6 (top). M- and Beta-value distribution before and after ComBat (bottom).*

Genetic matching: To confirm the absence of switched sample, we matched the SNP data provided from the Illumina EPICv2 array with Affymetrix-based genetic data from NESDA and compared the genotype. Doing so, we saw that out of 54 unique autosomal SNP probes present on the Illumina EPICv2 array, 31 provided a unreliable sample-independent signal. Of these probes, 13 overlapped with a list of 21 problematic probes from another recent study[9]. The remaining 23 SNP probes from Illumina EPICv2 had a consistent match to the Affymetrix data for all samples from all timepoints.

Cell-type proportion estimation: Based on raw .idat files deconvolution was performed by following the EPICv2 workaround given by the FlowSorted.Blood.EPIC package maintainers under <https://github.com/immunomethylomics/FlowSorted.Blood.EPIC/issues/10>. The pipeline returns six blood cell proportions: B cells, CD4+ T cells, CD8+ T cells, natural killer cells, monocytes and neutrophils. As the six proportions sum to one, one column is redundant. Monocytes were therefore dropped, and the remaining five (Bcell, CD4T, CD8T, NK, Neu) were used as covariates in all models.

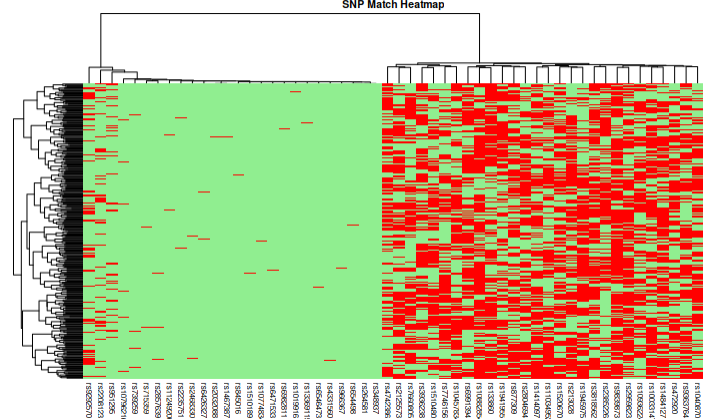

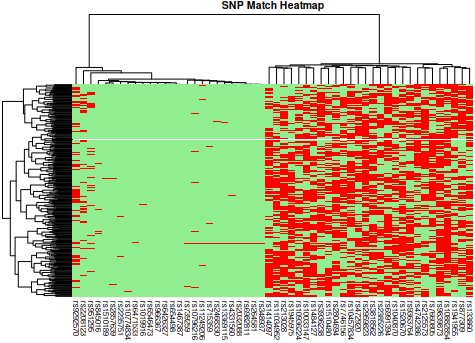

*Genetic matching based on SNP-probes from Illumina EPICv2 BeadChip for NESDA wave 1 (above) and wave 5 (below). Red signifies genotype mismatch, green signifies a match.*

Calculation of the methylation score for major depression (MD): The MD-MS is a weighted sum between EWAS-derived effect sizes and each of the epigenome-wide significant 15 CpG sites’ respective M value:

$$\text{MD-MS}_{j}=\sum_{i=1}^{15} \beta_{i} M_{ij}$$

where *j* indexes the individual sample, *i* indexes the CpG probe, *M*_ij_ the M-value for CpG *i* in individual *j* and *β*_i_ = the effect size for CpG *i* from the PGC EWAS meta-analysis for major depression. The outcome was scaled across all samples from both timepoints for linear mixed effects modelling. Among those CpG sites, only two (cg10604091, cg12805451) showed evidence for MD-related genetic effects in MR analyses performed by Shen et al., 2025 [10]:

|  | **Wald's ratio MR** | | | |
| --- | --- | --- | --- | --- |
|  | **Beta** | **SE** | **pval** | **pFDR** |
| cg10604091 | 0,932192 | 0,310731 | 0,0027 | 0,025558 |
| cg12805451 | 0,194256 | 0,071875 | 0,006878 | 0,042463 |

*Statistical analysis*

R Packages: The following packages were used: AnnotationHub v4.0.0, BiocParallel v1.44.0, broom v1.0.11, broom.mixed v0.2.9.6, cowplot v1.2.0, dplyr v1.1.4, effsize v0.8.1, emmeans v2.0.1, flextable v0.9.10, FlowSorted.Blood.EPIC v2.10.0, ggcorrplot v0.1.4.1, gghalves v0.1.4, ggplot2 v4.0.1, ggsankey v0.0.99999, ggsignif v0.6.4, gt v1.3.0, gtsummary v2.5.0, haven v2.5.5, IlluminaHumanMethylation450kanno.ilmn12.hg19 v0.6.1, IlluminaHumanMethylationEPICanno.ilm10b4.hg19 v0.6.0, IlluminaHumanMethylationEPICv2anno.20a1.hg38 v1.0.1, inspectdf v0.0.12.1, lme4 v2.0.1, lmerTest v3.2.0, minfi v1.56.0, officer v0.7.3, patchwork v1.3.2, pheatmap v1.0.13, polycor v0.8.2, pROC v1.19.0.1, purrr v1.2.2, rstatix v0.7.3, rtracklayer v1.70.1, sesame v1.28.1, sva v3.58.0, tidyr v1.3.2, writexl v1.5.4.

Effect size and AUC of MD-MS: The association between lifetime MDD (and the other categorical clinical phenotypes listed in **Table 2**) and the methylation score was estimated across both visits in a linear mixed model:

$$\text{MD-MS}=\beta_{0}+\beta_{1}\text{MDD}+\beta_{2}\text{Sex}+\beta_{3}\text{Age}+\beta_{4}\text{Age}^{2}+\beta_{5}\text{Bcell}+\beta_{6}\text{CD4T}+\beta_{7}\text{CD8T}+\beta_{8}\text{NK}+\beta_{9}\text{Neu}+\left( 1 | \text{Subject ID} \right)+ \varepsilon$$

where MD-MS is the scaled methylation score for major depression, Bcell, CD4T, CD8T, NK and Neu are the scaled blood cell-type proportions, and (1|Subject ID) is a participant-level random intercept accounting for the correlation between a participant’s paired observations. Age was entered as a second-degree polynomial (poly(age, 2)), yielding uncorrelated linear and quadratic terms. Models were fitted with the lmer() function from the lmerTest package, which supplies Satterthwaite-approximated degrees of freedom for the fixed-effect *t*-tests. The regression coefficient, standard error, 95% confidence interval and *P*-value for β₁ were extracted with tidy(effects = "fixed", conf.int = TRUE) from the broom.mixed package. With lifetime MDD coded FALSE/TRUE, β₁ is the adjusted mean difference in MD-MS between lifetime cases and lifetime healthy participants, in score standard deviations. To derive the effect size, β₁ was standardized as a Cohen's *d*-analogue relative to the total residual standard deviation:

$$d\mathbf{=}\frac{\beta_{\mathbf{1}}}{\sqrt{\tau^{\mathbf{2}}\mathbf{+}\sigma^{\mathbf{2}}}}$$

where τ² is the between-person (random-intercept) variance, obtained from VarCorr() from the lme4 package, and σ² the residual variance, obtained from sigma() from base R. We computed *d* with the eff_size() function from the emmeans package. The “sigma” argument was set to the total standard deviation above, and “edf” was set to the Satterthwaite degrees of freedom. For continuous predictors listed in Table 2, the predictor was z-scored and the standardised slope obtained with emtrends() was divided by the same denominator, with confidence limits from the *t*-distribution with Satterthwaite degrees of freedom.

To quantify how well the methylation score discriminates lifetime cases from lifetime healthy participants, the area under the receiver operating characteristic curve (AUC) was computed. Since the score is influenced by age, sex and blood cell composition, the raw score would confound discrimination by MDD with discrimination by these covariates; the score was therefore first residualised against a covariate-only model fitted in the same sample:

$$MD-MS=\beta_{0}+\beta_{1}Sex+\beta_{2}Age+\beta_{3}\mathrm{Age}^{2}+\beta_{4}Bcell+\beta_{5}CD4T+\beta_{6}CD8T+\beta_{7}NK+\beta_{8}Neu+\left( 1 | \mathrm{pident} \right) + \varepsilon$$

The residualised score was obtained by subtracting the marginal prediction of this model – that is, the prediction from the fixed effects alone, denoted fixed:

$$\text{MD-MS}_{\text{resid}}=\text{MD-MS}-\hat{y}_{fixed}$$

Marginal predictions were generated with predict() setting the argument “re.form” to NA, which instructs lme4 to exclude the random-intercept terms from the prediction to retain between-person variance in the residual. The AUC of the residualised score against lifetime MDD status was then computed with the roc() function from the pROC package, taking the binary outcome and the residualised score as inputs, with “ci” set to TRUE. The point estimate and its 95% confidence interval were extracted with pROC functions auc() and ci.auc(), which uses DeLong's method for a single ROC curve.

Marginal residualisation was chosen as the primary approach because the random intercept captures stable between-person variance in the score, which includes the variance associated with lifetime MDD, a largely time-invariant person-level characteristic. Because the covariate-only model does not include diagnosis, removing the random intercept (conditional residualisation) removes part of the signal of interest. The AUC based on conditional residuals, which removes the person-specific random intercept and leaves predominantly within-person variation, was closer to chance (AUC = 0.54 [0.497, 0.582]). This suggests that the discriminative information in the score resides in the stable between-person component and not in visit-to-visit variation.

Linear mixed effect model specifications:

Covariates in all cross-sectional models were age, age^2^, sex, cell type proportions (DNAm-estimated B-cells, CD4^+^ T-cells, CD8^+^ T-cells, natural killer cells and neutrophils [11]). In the longitudinal models, age/age^2^ was replaced by baseline age/age^2^; visit (baseline vs. FU6) and diagnostic group (lifetime healthy, new onset, remitted, chronic MDD) were added and cell type proportions were person-mean decomposed into between- and within-person terms.

Cross-sectional analyses shown in Figure 1 and Figure 2 have been derived from linear mixed effect models with random intercepts for pooled paired samples from baseline and FU6 using the lmerTest package. We fitted three cross-sectional model types: univariate models including lifetime MDD (or other clinical predictors) as variable of interest (Figure 1, Table 2), univariate models including the domain variables of interest (psychosocial, environment, biological, lifestyle, Figure 2) and multivariate models including both lifetime MDD and domain variables (Figure 2). In order to justly compare the reduction of lifetime MDD’s regression coefficient from univariate to multivariate model, we fitted a univariate model for lifetime MDD including only those samples from the multivariate model with complete data

The longitudinal association of MD-MS with the four groups capturing MDD clinical trajectories (new onset, chronic, remitted MDD, and healthy) were examined in separate linear mixed-effects models. Visit-by-group interaction terms were later included to test whether MD-MS trajectories differed across groups over time with reference to the lifetime healthy group. In a secondary analysis, as alternative to the four MDD trajectories, we decomposed the IDS score at baseline and FU6 into its between- and within-person components via the person-mean centering approach; the visit term was kept in the model formula. All numerical variables were scaled using the base R *scale()* function. Estimated marginal means were derived using the *emmeans* package to visualize group-specific trajectories.

*Used models*

**Model 1 – Lifetime MDD / clinical predictor (Figure 1, Table 2)**

MD-MS ~ Predictor + Age + Age² + Sex + Cells + (1 | subject)

*Predictor = lifetime MDD, IDS, antidepressant use, current anxiety, BAI, age of onset, chronicity, or lifetime anxiety (one per model)*

**Model 2 – Single domain variable (Figure 2)**

MD-MS ~ DomainVariable + Age + Age² + Sex + Cells + (1 | subject)

**Model 3 – Lifetime MDD + all variables of one domain (Figure 2)**

MD-MS ~ MDD + DomainVariable_1 + … + DomainVariable_M + Age + Age² + Sex + Cells + (1 | subject)

Model 1 (Predictor = lifetime MDD) is refitted on the complete-case sample of Model 3.
% reduction = (b_MDD,Model1 − b_MDD,Model3) / |b_MDD,Model1| × 100

**Model 4 – MDD trajectories (Figure 3)**

MD-MS ~ Visit + Group + Visit + Group + Age_baseline + Age_baseline² + Sex + Cells_between + Cells_within + (1 | subject)

MD-MS ~ Visit + Group + Visit × Group + Age_baseline + Age_baseline² + Sex + Cells_between + Cells_within + (1 | subject)

Group = new onset, remitted, chronic (reference: lifetime healthy).

**Model 5 – Depressive symptoms (Table 3)**

MD-MS ~ Visit + IDS_between + IDS_within + Age_baseline + Age_baseline² + Sex + Cells_between + Cells_within + (1 | subject)
